# Simultaneous whole-head electrophysiological recordings using EEG and OPM-MEG

**DOI:** 10.1101/2023.10.22.23297153

**Authors:** Zelekha A. Seedat, Kelly St Pier, Niall Holmes, Molly Rea, Layla Al-Hilaly, Tim M. Tierney, Rosemarie Pardington, Karen J. Mullinger, J. Helen Cross, Elena Boto, Matthew J. Brookes

## Abstract

Electroencephalography (EEG) and magnetoencephalography (MEG) non-invasively measure human brain electrophysiology. They differ in nature; MEG offers better performance (higher spatial precision) whilst EEG (a wearable platform) is more practical. They are also complementary, with studies showing that concurrent MEG/EEG provides advantages over either modality alone, and consequently clinical guidelines for MEG in epilepsy recommend simultaneous acquisition of EEG. In recent years, new instrumentation – optically pumped magnetometers (OPMs) – has had a significant impact on MEG, offering improved performance, lifespan compliance, and wearable sensors. Nevertheless, the ability to carry out simultaneous EEG/OPM-MEG remains critical. Here, we investigated whether simultaneous, wearable, whole-head EEG and OPM-MEG measurably degrades signal quality in either modality. We employed two tasks: a motor task known to modulate beta oscillations, and an eyes-open/eyes-closed task known to modulate alpha oscillations. In both, we characterised the performance of EEG alone, MEG alone, and concurrent EEG/OPM-MEG. Our results show that the SNR of the beta response was very similar, regardless of whether modalities were used individually or concurrently. Likewise, our alpha band recordings demonstrated that signal contrast was stable, regardless of the concurrent recording. These results combined suggest that there are no fundamental barriers to simultaneous wearable EEG/OPM-MEG, and consequently this technique is ripe for neuroscientific and clinical adoption. This will be particularly important in the clinical sphere where a direct comparison between simultaneous EEG and OPM-MEG recordings will facilitate interpretation of OPM-MEG data in patients.

## INTRODUCTION

Electroencephalography (EEG) (Berger, 1929) and magnetoencephalography (MEG) (Cohen, 1968) provide non-invasive characterisation of brain electrophysiology. EEG records changing electrical potential difference at the scalp caused by current flow through neural assemblies. MEG measures changing magnetic fields above the scalp generated by similar currents. Both modalities allow interrogation of brain function in health and disease and offer significant clinical utility – particularly in disorders like epilepsy where a change in the electrical activity underlies symptoms.

EEG and MEG differ in both practicality and performance (Baillet, 2017): EEG is ‘wearable’ (meaning lightweight sensors are placed in electrical contact with the scalp and move freely with the head, allowing adaptability to any head shape/size and free movement during data acquisition). This allows it to be well tolerated and lifespan compliant. It is also readily available and low cost. However, the high electrical resistance of the skull reduces the amplitude of measured potentials and spatially distorts the signal topography, meaning sensitivity and spatial resolution are limited. EEG is also susceptible to interference from muscles (Whitham, et al., 2007) which degrades data quality. Conversely, MEG is less sensitive to muscle artifacts (Muthukumaraswamy, 2013) (Boto, et al., 2019) and offers better spatial resolution than EEG (since magnetic fields pass through the skull with little distortion). However, conventional MEG systems rely on superconducting sensors to measure the neuromagnetic field (Hamalainen, Hari, Ilmoniemi, Knuutila, & Lounasmaa, 1993). Because such sensors require low temperatures, they are fixed in a rigid helmet with a gap between the sensors and the scalp for thermal insulation. Consequently, signal magnitude is limited by sensor proximity; a problem that is more prominent in participants with small heads. Moreover, participants must remain still relative to the fixed sensor array, which makes MEG hard to deploy in many participants (e.g., infants). In summary, whilst MEG offers improved performance, EEG has significant practical advantages due to its wearability.

Despite their differences, a large body of evidence shows that EEG and MEG are complementary. MEG is most sensitive to current flow oriented tangential to the scalp whereas EEG is most sensitive to radial currents. This means, theoretically, simultaneous measurement offers improved coverage. This is realised in practice; for example, Aydin et al. (Aydin, et al., 2015) showed that, in epilepsy, simultaneous EEG/MEG allowed mapping of spike propagation that was not possible via MEG or EEG alone. Likewise Yoshinaga et al (Yoshinaga, et al., 2002) used dipole modelling to show that MEG and EEG provides a level of information that was not obtainable with either modality alone. These demonstrations show the importance of combining the two methods, and clinical guidelines reflect this by recommending concurrent recording in epilepsy (Bagic, Knowlton, Rose, & Ebersole, 2011). However, the fixed nature of MEG means that, in concurrent EEG/MEG, the significant practical advantages of EEG are lost, and participants must remain still in a cumbersome machine, ruling out naturalistic behaviour and reducing patient comfort.

In recent years MEG has been revolutionised by the introduction of new magnetic field sensors (see e.g., (Tierney, et al., 2019) (Brookes, et al., 2022) (Schofield, et al., 2023) for reviews). Optically Pumped Magnetometers (OPMs) are small and lightweight sensors which measure magnetic field with a sensitivity comparable to superconducting sensors, but without cryogenic cooling. Because OPMs can get closer to the scalp than cryogenic sensors, they detect a signal that is less spatially diffuse and higher amplitude, bringing improved sensitivity and spatial resolution (Boto, et al., 2016) (Borna, et al., 2017) (Iivanainen, Stenroos, & Parkkonen, 2017) (Nugent, Andonegui, Holroyd, & Robinson, 2022). Moreover, OPMs can be mounted in a lightweight helmet and move with the head, meaning that (assuming background field is controlled or a closed loop system is employed (Holmes, et al., 2018) (Holmes, et al., 2019) (Mellor, et al., 2022) (Robinson, et al., 2022)) the MEG signal can be measured as a person moves (Boto, et al., 2018). Whole head OPM-MEG systems are emerging (e.g., (Hill, et al., 2020) (Boto, et al., 2021) (Rea, et al., 2022) (Alem, et al., 2023) (Rhodes, et al., 2023)) and the clinical potential of OPM-MEG is also being shown (Vivekananda, et al., 2020) – for example OPM-MEG offers higher sensitivity (compared to conventional MEG) for detection of epileptic spikes in children (Feys, et al., 2022); it can record patient data during a seizure (Feys, et al., 2022) (Hillebrand, et al., 2023) and a recent case study reported that, even for a deep source (mesiotemporal cortex), it could measure ∼60% of the epileptic discharges that were identified using invasive EEG (Feys, et al., In Submission). These initial studies indicate that OPM-MEG therefore provides the technical advantages of MEG within a package that is similar in form to EEG – a wearable, motion robust helmet.

OPM-MEG now opens the opportunity for the combined use of EEG and MEG within a single wearable system. Such a system would adapt to head shape/size and allow free head movement during data acquisition. It would enable the additional information content from concurrent MEG/EEG recordings without the loss of the major advantages of EEG. Perhaps most significantly, simultaneous measurements would allow clinicians to reap the significant advantages that OPM-MEG brings (over conventional MEG and EEG) whilst maintaining the clinical standard (EEG). Two previous studies (Boto, et al., 2019) (Xingyu, et al., 2022) already show that OPMs can operate in close proximity to EEG electrodes. However, Boto et al. had only two OPMs and it was not possible at that stage to attain whole head coverage. Similarly, Xingyu et al. used an integrated MEG-EEG-fNIRS system, but OPMs were placed over right frontal and temporal lobes only (not whole-head coverage) and there was no assessment of the impact on MEG, EEG or fNIRS data quality in the presence of the two other recording modalities. Here, we expand on these studies by demonstrating the simultaneous use of whole head (64 channel) EEG and whole head (64 dual-axis sensor, 128 channel) OPM-MEG. In this study we collected data in 12 individuals using 1) EEG alone 2) OPM-MEG alone and 3) concurrent EEG/OPM-MEG. We restrict ourselves to channel level analyses (to avoid confounding effects due to e.g., differences in source localisation strategies) and we quantitatively compute signal to noise ratio (SNR) to test the hypotheses that: 1) The presence of whole head OPM-MEG does not significantly degrade the SNR of EEG recordings and 2) the presence of whole head EEG does not significantly degrade the SNR of OPM-MEG recordings. Successful confirmation of these hypotheses would demonstrate the utility of concurrent EEG/OPM-MEG for future clinical (and basic science) applications.

## MATERIALS AND METHODS

This research was approved by the University of Nottingham Medical School Research Ethics Committee. Study participants were 12 adults aged between 26 and 64 (mean age 41, 8 female) with no known neurological conditions. They all gave written informed consent for the study. The data and code used in this study are available on GitHub: https://github.com/ZSeedat/EEG_OPMMEG_HealthyAdultStudy_YoungEpilepsy

### Hardware

Data were acquired at Young Epilepsy (Lingfield, Surrey, UK). The OPM-MEG device is a complete integrated system (Cerca Magnetics Ltd. Nottingham, UK) including a sensor array, helmet for sensor mounting, patient support, magnetic shielding and field control. It also includes equipment for participant motion tracking, coregistration of sensor locations to brain anatomy, stimulus delivery and data acquisition (including delineation of stimulus timing).

The OPM array comprised 64 dual-axis magnetic field sensors (QuSpin Inc, Colorado, USA) which provide 128 independent measurements of field around the head (128 channels). The sensors are integrated into a single system to simultaneously measure field and can be mounted in a series of rigid helmets of varying size, to accommodate individuals of any age.

To achieve a magnetically “quiet” environment, the system is housed in a magnetically-shielded room (MSR) (Holmes, et al., 2022) comprising multiple layers of high permeability / conductivity metal which reduce both static and time-varying magnetic fields. The room is equipped with degaussing coils and these were used to demagnetise the innermost metal layer, which results in a reduction of static field beyond that afforded by the room itself. The room is also equipped with a set of 27 “window” coils embedded within the walls. Given a measured field in the room, these coils can be independently controlled such that they generate a summed magnetic field which opposes the measured field, enabling precise field nulling in the space occupied by the participant. This in turn ensures minimal magnetic artifact as a participant moves.

The system includes a PC for delivering stimuli to the participant and instrumentation to facilitate time-locking between the stimulus and measured data (i.e. a “triggering” system). It has cameras for tracking participant motion (OptiTrack, NaturalPoint Inc., Oregon, USA) (via monitoring movement of infrared retro-reflective markers) and equipment for coregistration of sensor positions to brain anatomy.

A 64-channel MEG-compatible EEG system (Brain Products GmbH, Munich, Germany) was used to acquire all EEG data. This system comprised an EEG-cap (with passive, MEG compatible, Ag/AgCl electrodes), signal amplifiers, a power pack, a USB adaptor box and a data acquisition laptop. The ground electrode was AFz and the reference electrode was FCz. 63 channels of data were recorded from the scalp, the 64^th^ channel recorded the electrocardiogram (ECG). EEG electrodes were connected to the head with a conductive gel, and the impedances of all good electrodes were kept below 10kΩ on all subjects. Triggers from the stimulus PC were split so that simultaneous markers appeared in the OPM-MEG and EEG data. This enabled synchronisation of the OPM-MEG and EEG data as well as data from peripheral recordings (e.g., motion tracking). Participants sat in a patient support at the centre of the MSR and a two-way intercom enabled communication between the experimenter and the participant. Figure 1A shows a schematic of the system used. Figure 1B is an outside view of the MSR, and Figure 1C is a photo of the EEG and MEG helmets in place on a participant.

**Figure 1:**
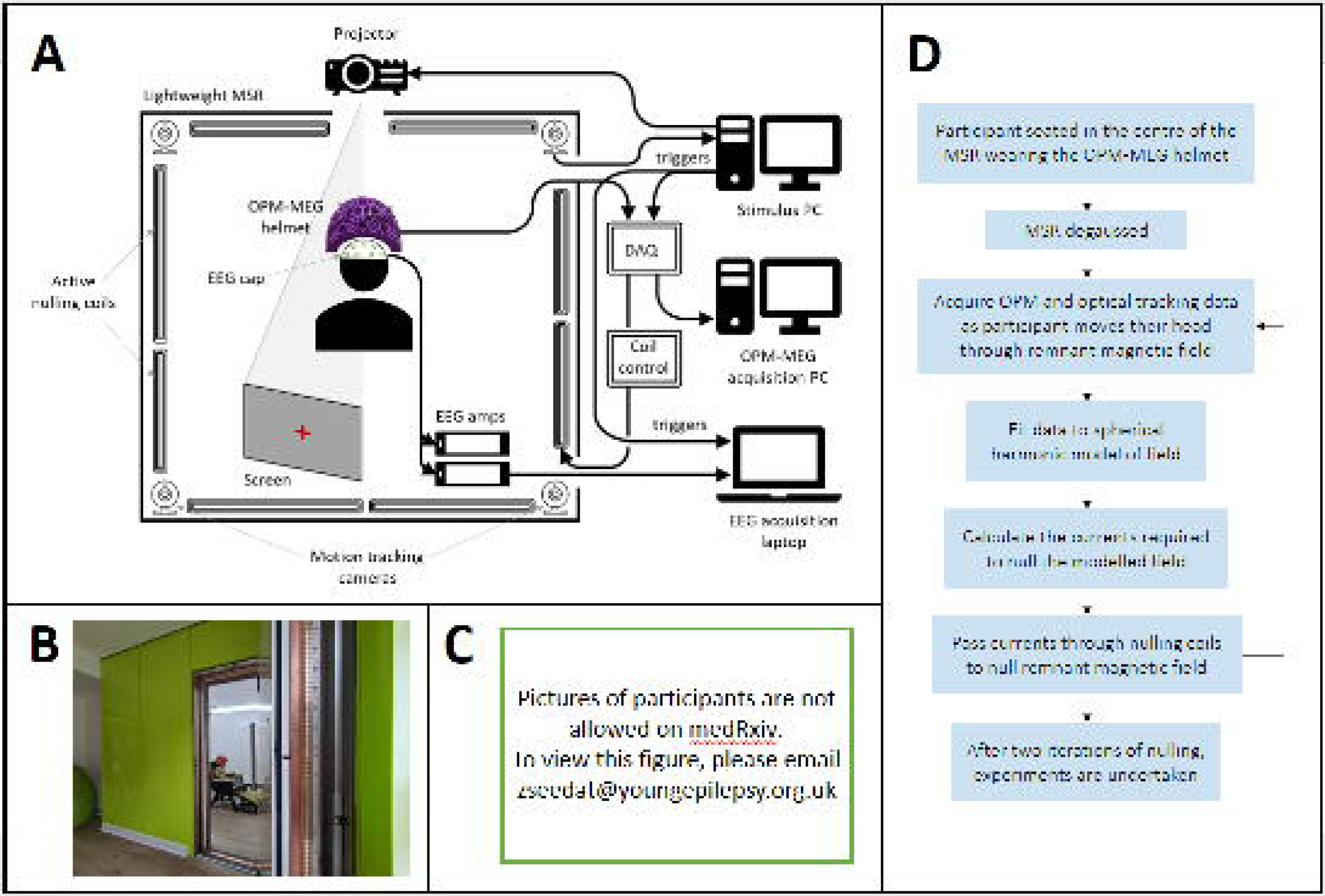
Experimental set-up and field nulling. (A) Schematic of the simultaneous OPM-MEG/EEG system. (B) The lightweight magnetically shielded room at Young Epilepsy where experiments took place. (C) Participants wore the OPM-MEG helmet (with or without an EEG cap). (D) Flow chart describing the field nulling process.

### Experimental paradigms

Each participant underwent 3 experimental sessions: OPM-MEG only; EEG only and simultaneous EEG/OPM-MEG. Each session took place in the MSR and lasted ∼40 mins. The order of sessions and tasks was pseudorandomised across participants and all sessions were completed on the same day. Within each session, participants performed 5 tasks, however only 2 are reported here:

1. **Motor task:** Participants wore a motion tracking marker on their right index finger and were asked to complete one brief finger abduction when a visual cue (an image of a hand) appeared on the screen. A single trial was 5 s in duration, and the visual cue was presented for 2 s. In the rest phase a red fixation cross was shown. A total of 50 trials was recorded. This task elicits a robust change in beta (13-30Hz) amplitude, including a movement-related beta decrease (MRBD) and a post-movement beta rebound (PMBR) (Pfurtscheller & Lopes da Silva, 1999).
2. **Alpha generation task:** Participants were asked to look at a red fixation cross on a grey screen. Every 30s they were asked by the experimenter to either close or open their eyes. This was repeated 5 times (i.e. 5 trials, each with duration 1 minute). This is well known to drive increases or decreases in occipital alpha oscillations (Berger, 1929).

### Nulling background fields

OPMs are vector field sensors, meaning if they move relative to a non-zero background field they measure a signal that could obfuscate the neural signals of interest, and potentially stop the OPMs from working (Holmes, et al., 2018). For this reason, nulling the field inside the MSR is important. Here, we used an approach originally described by Rea et al (Rea, et al., 2021). Briefly, the helmet was positioned on the participant’s head, the MSR door closed and the room degaussed. The participant was then asked to execute a series of simple head movements. During these movements we simultaneously recorded the magnetic field variations at the OPMs, and head motion (via the motion tracking cameras). These synchronised data were input into a fitting algorithm to characterise the remnant field and its first order gradient across the volume occupied by the helmet. The resulting field was used to determine coil currents which were then applied to the window coils. This generated a field equal and opposite to that measured, thereby cancelling it out. This procedure was carried out twice for each participant, followed by a third field map to measure the final fields in the room. This is described in Figure 1D.

### Data acquisition

Field nulling – as described above – was carried out for experimental sessions where OPM-MEG was used (not for sessions where EEG was used alone). For all experiments and sessions, OPM-MEG and EEG data were recorded at 1200Hz and 1000Hz respectively. Visual stimuli were shown via projection through a waveguide behind the participant onto the wall of the MSR (visual angle 42° horizontally and 27° vertically). Motion tracking data (to monitor the finger abduction in the motor task) was captured simultaneously with OPM-MEG and EEG at a sample rate of 120Hz; synchronisation was achieved by simultaneous triggers, sent from the stimulus PC to all three recordings (MEG, EEG and motion tracking).

### Preprocessing

Pre-processing was performed in MATLAB using the FieldTrip toolbox (Oostenveld, Fries, Maris, & Schoffelen, 2011) and the same pipeline was used for EEG and OPM-MEG. Data were bandpass filtered between 1 and 150Hz using a 4^th^ order Butterworth filter, notch filtered at 50Hz to remove mains frequency noise (also 4^th^ order Butterworth), baseline corrected, and detrended with a linear fit. Data were then segmented into trials for each task:

- Motor task trials were defined as 1 s before movement cessation (measured by the motion tracking cameras) to 4s after movement cessation (5-s duration).
- Alpha trials were defined from 1 s after the instruction to open/close their eyes (to allow for slower reaction times) until 6 s after the instruction (i.e., 5-s trial duration). This time window was taken because participants who are alert with their eyes closed are known to elicit the highest amplitude alpha response (Nunez, 2002) and towards the end of the eyes closed period they are likely to be in a drowsy state which would attenuate alpha.

The signal variance for each trial and channel was inspected visually, and outliers (i.e. those trials with obvious artifacts) were removed. Homogeneous field correction (to reduce any effect of external interference) was applied to OPM-MEG data (Tierney, et al., 2021) and EEG data were re-referenced (to the average of all good channels for the motor task, and the average of all good channels excluding those over occipital areas (namely: Pz, POz, Oz, P1-8, PO3, PO4, PO7-10, O1, O2) for the alpha task). To ensure that our results were robust to changes in the EEG referencing scheme, all EEG analyses were repeated using the common recording reference (CRR) and these results are shown in the supplementary information.

For the motor task, 2 participants were excluded from further analyses because there was no task-modulated response in EEG or MEG. One further participant was excluded due to missing data (motion tracking and the EEG-MEG synchronisation trigger failed), this left 9 participants. For the alpha generation task, no participants were excluded. Tables 1 and 2 show the numbers of trials and channels left after outliers had been removed.

**Table 1:** The numbers of participants, good channels, and good trials after pre-processing for the motor task.

| Motor Task | EEG only | MEG only | EEG with MEG | MEG with EEG |
| --- | --- | --- | --- | --- |
| No. of participants | 9 | 9 | 9 | 9 |
| No. of good channels<br>(mean ± std) | $57 \pm 2$ | $127.4 \pm 0.9$ | $57 \pm 2$ | $126 \pm 1$ |
| No. of good trials<br>(mean ± std) | $48 \pm 4$ | $47 \pm 2$ | $48 \pm 2$ | $47 \pm 2$ |

**Table 2:** The numbers of participants, good channels, and good trials after pre-processing for the alpha generation task.

| <u>Alpha Generation</u> | EEG only | MEG only | EEG with MEG | MEG with EEG |
| --- | --- | --- | --- | --- |
| No. of participants | 12 | 12 | 12 | 12 |
| No. of good channels<br>(mean ± std) | 59 ± 2 | 126 ± 2 | 58 ± 2 | 126 ± 1 |
| No. of good trials<br>(mean ± std) | 5 ± 0 | 4.9 ± 0.3 | 5 ± 0 | 4.8 ± 0.4 |

### Field nulling analysis

To estimate the efficacy of the field nulling, the root mean square (RMS) magnetic field from the uniform field and field gradients components was computed on a spherical surface containing the head, and the range of head movements. A sphere of radius 0.25 m was chosen based on the range of head movements during field mapping.

### Motor task analysis

Following pre-processing, time-frequency spectrograms (TFSs) were computed for every OPM-MEG and EEG channel: briefly, data were band-pass filtered into 18 overlapping frequency bands using a series of 3^rd^ order Butterworth filters. The signal from each band was Hilbert transformed to compute the analytic signal, and the absolute value taken to give the envelope of oscillatory amplitude. Concatenation in the frequency dimension produced the TFS, which was averaged across trials. The TFS was baseline corrected by subtracting the mean amplitude in the 3-4s time window (relative to movement offset). The TFS was then divided by the baseline, to give a measure of relative change.

For all channels (OPM-MEG and EEG) we estimated beta band signal to noise ratio (SNR). An MRBD window was defined as -1 s to 0 s, and a PMBR window defined as 0.5 s to 1.5 s (both relative to movement cessation). SNR was calculated as the difference in mean signal amplitude between the two windows, divided by the standard deviation of the signal in the MRBD window. SNR was calculated for all channels, and data from channels with the highest SNR were averaged across participants. We tested for statistically significant differences: i.e., if OPM-MEG SNR was changed by the presence of EEG, and if EEG SNR was changed by OPM-MEG. This was assessed using a Wilcoxon rank-sum test.

In addition to SNR, we derived a measure of how far the signal from motor cortex spreads across scalp electrodes/sensors (as a result of volume conduction (EEG) or magnetic field propagation (OPM-MEG)). For both EEG and OPM-MEG data, we measured Pearson correlation between the signal from the channel with the highest SNR, and all other channels (signals were beta envelopes, unaveraged across trials). We then counted the number of channels with a correlation above 0.3 and designated them “highly correlated”. The proportion of highly correlated channels was then calculated for each participant and averaged. This was carried out independently for EEG and OPM-MEG.

### Alpha generation task analyses

Data from the alpha task were initially processed using independent component analysis (ICA) to remove the heartbeat artifact (using the AnyWave (Colombet, Woodman, Badier, & Bénar, 2015) infomax implementation of ICA). Data were then bandpass filtered between 2 and 40Hz, and notch filtered at 50Hz (again using AnyWave). The filtered EEG and OPM-MEG data were inspected visually to look for the presence of alpha oscillations. Spectral analysis was also undertaken: data were frequency filtered in the 1-150Hz band using a 4^th^ order Butterworth filter. Data were then segmented into trials and power spectral density (PSD) in the eyes open and eyes closed windows was estimated using the MATLAB periodogram. We then computed a measure of signal contrast, defined as 8-13Hz spectral power with eyes closed, divided by eyes open. The channel with the highest contrast was selected for each participant and we averaged PSDs across individuals. We then tested for differences in contrast between sessions with or without simultaneous recordings.

## RESULTS

### Nulling background fields

Field nulling experiments were completed successfully in 11/12 participants, with technical difficulty on the day of scanning preventing measurement in one person. Results are shown in Figure 2; before nulling, the RMS field over a spherical volume, with and without EEG, was 4.5nT ± 0.6nT and 5.0nT ± 0.5nT (mean ± standard deviation across participants) respectively. This dropped to 0.7nT ± 0.5nT and 0.7nT ± 0.3nT (with and without EEG, respectively) after field nulling. A Wilcoxon rank-sum test suggested no significant difference in the efficacy of nulling (p = 0.5), with and without EEG.

**Figure 2:**
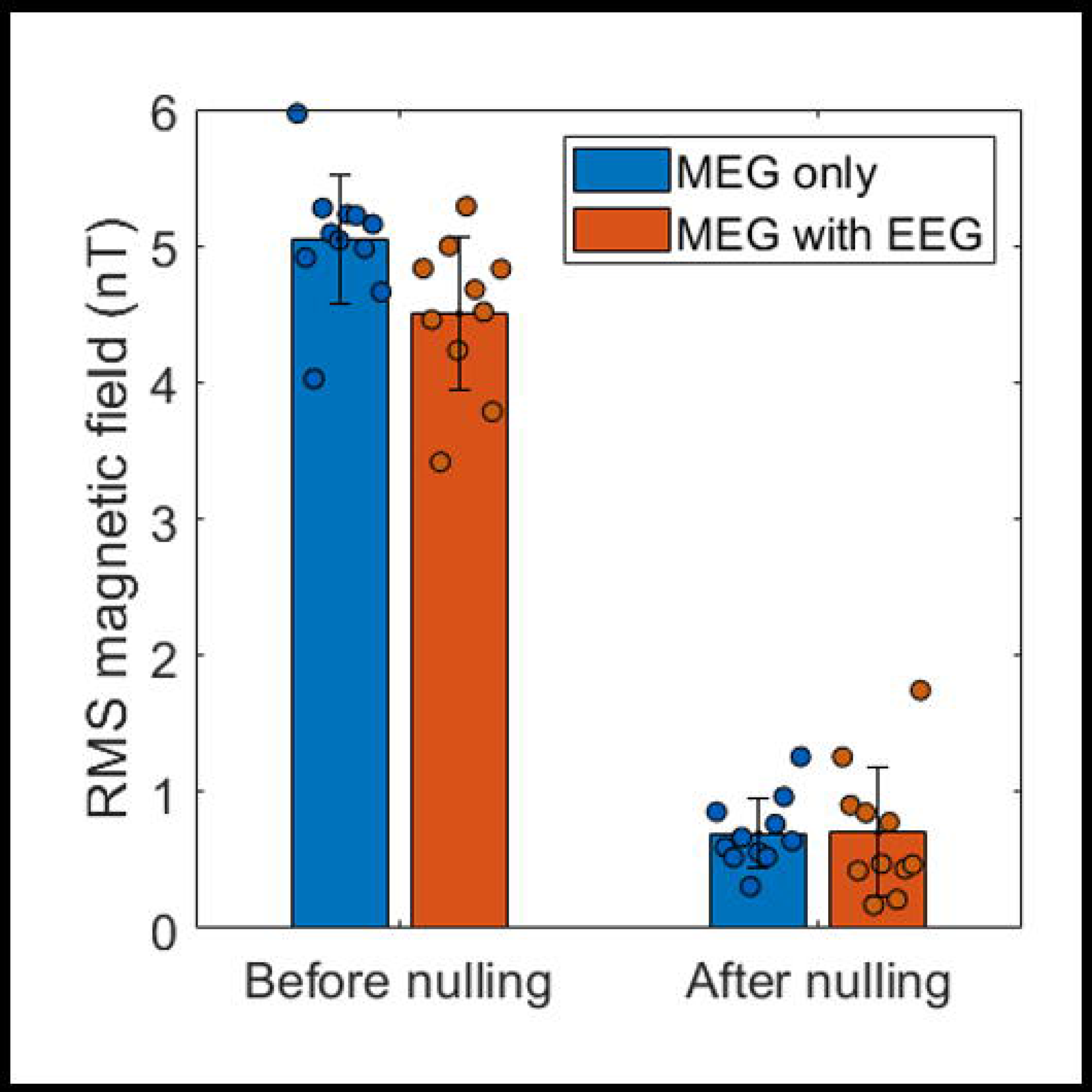
The presence of EEG does not significantly affect the efficacy of the field nulling process. Scatter points show the RMS field for individual sessions, bars show the mean, and error bars show the standard deviation. Blue bars show the RMS field before and after nulling for MEG only, and the orange bars are for MEG in the presence of EEG.

### Motor Task

Table 1 shows the number of participants, trials, and channels remaining in each dataset after preprocessing. Figure 3, panels A-D, show TFS’s and beta band oscillatory envelopes for the finger abduction task. In all cases, data have been averaged across participants and, in the case of the line plots, the shaded area represents standard deviation across participants. EEG data, with and without OPM-MEG, are shown in panels A and B respectively. OPM-MEG data, with and without EEG, are shown in panels C and D.

**Figure 3:**
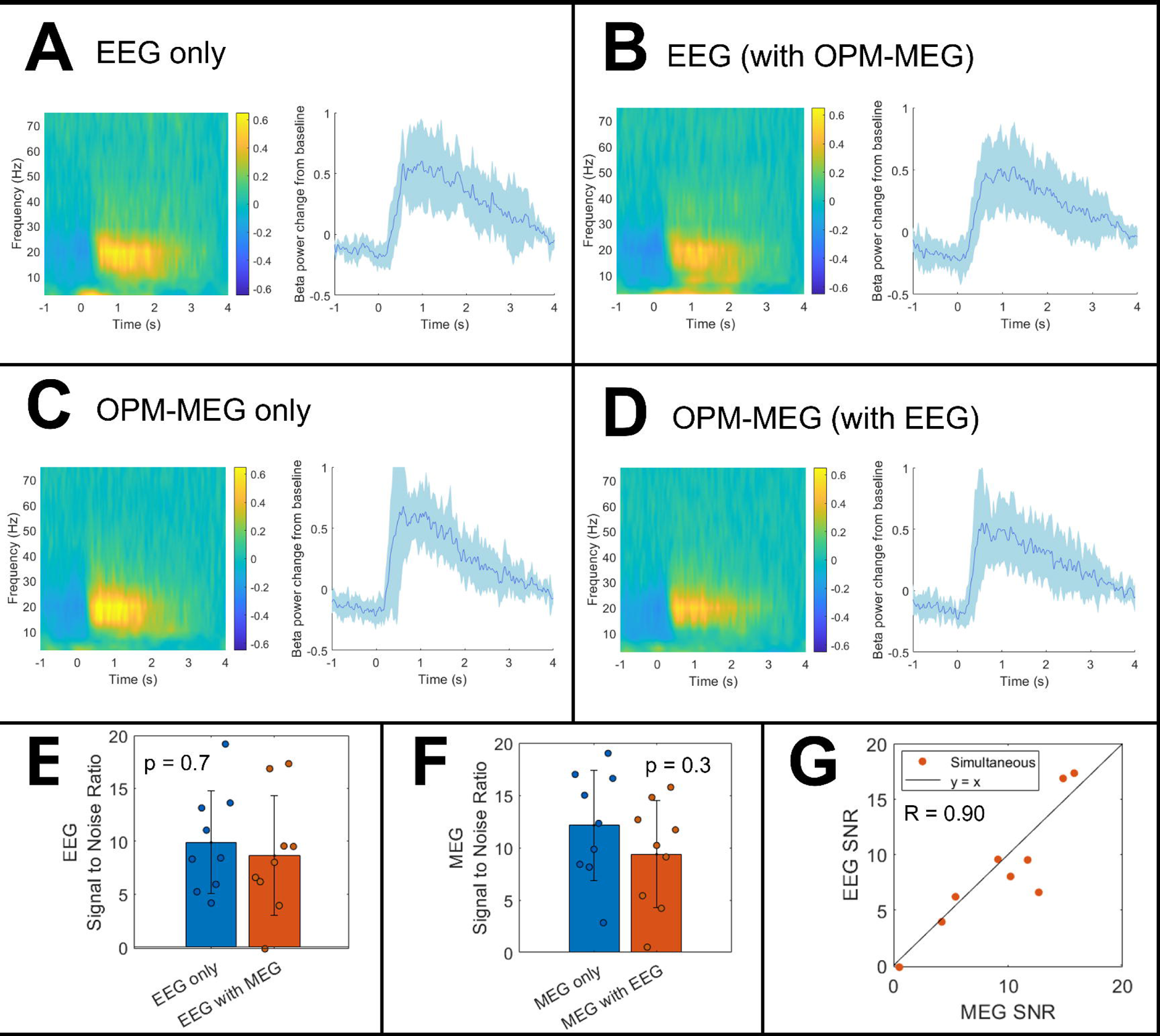
Finger abduction results. Panels A and B show the average TFS (left) and beta band envelope (right) in the peak channel for EEG alone and EEG in the presence of OPM-MEG, respectively. In both plots, a time of 0 s indicates the offset of finger movement. Panels C and D show equivalent responses for OPM-MEG only and OPM-MEG in the presence of EEG, respectively. Panel E shows SNR values for EEG, and Panel F shows equivalent values of OPM-MEG. Each data-point represents a participant; the bars represent the mean over participants, and the error bar describe standard deviation. Panel G plots EEG SNR against OPM-MEG SNR, for the simultaneously recorded data; again, a single data point represents an individual participant, and the black solid line represents y=x.

In all cases, the MRBD and PMBR are clearly visible and, most importantly, the addition of the simultaneous recording appears to have little effect on the result. SNR values for the peak channels are shown in Figures 3E (for EEG) and 3F (for OPM-MEG). Considering EEG, the SNR when used alone was 10 ± 5 (mean ± std) and dropped marginally to 9 ± 6 when OPM-MEG was added, but this was not significant (p = 0.7; Wilcoxon rank-sum test). For OPM-MEG, when used alone the SNR was 12 ± 5 which dropped to 9 ± 5 when the EEG was added, but again this was a non-significant effect (p = 0.3; Wilcoxon rank-sum test). Figure 3G shows the SNR of OPM-MEG plotted against the SNR of EEG for simultaneously acquired data; as expected, participants with low SNR in OPM-MEG also have low SNRs in EEG (Pearson correlation 0.9; p = 0.7).

In Figure 4, panels A and B show TFS data as a function of channel location for a single representative participant (for OPM-MEG, only the radially oriented channels are shown for ease of visualisation). Note that, qualitatively, the signal from the motor cortex appears to impact more scalp locations in EEG than it does for OPM-MEG. This is formalised in Figure 4C where the proportion of channels that are highly correlated with the peak response is shown for the two modalities. A Wilcoxon rank-sum test showed that there were significantly fewer highly correlated channels (p = 4 x 10^-5^) in OPM-MEG compared to EEG. This is an important finding and will be discussed further below.

**Figure 4:**
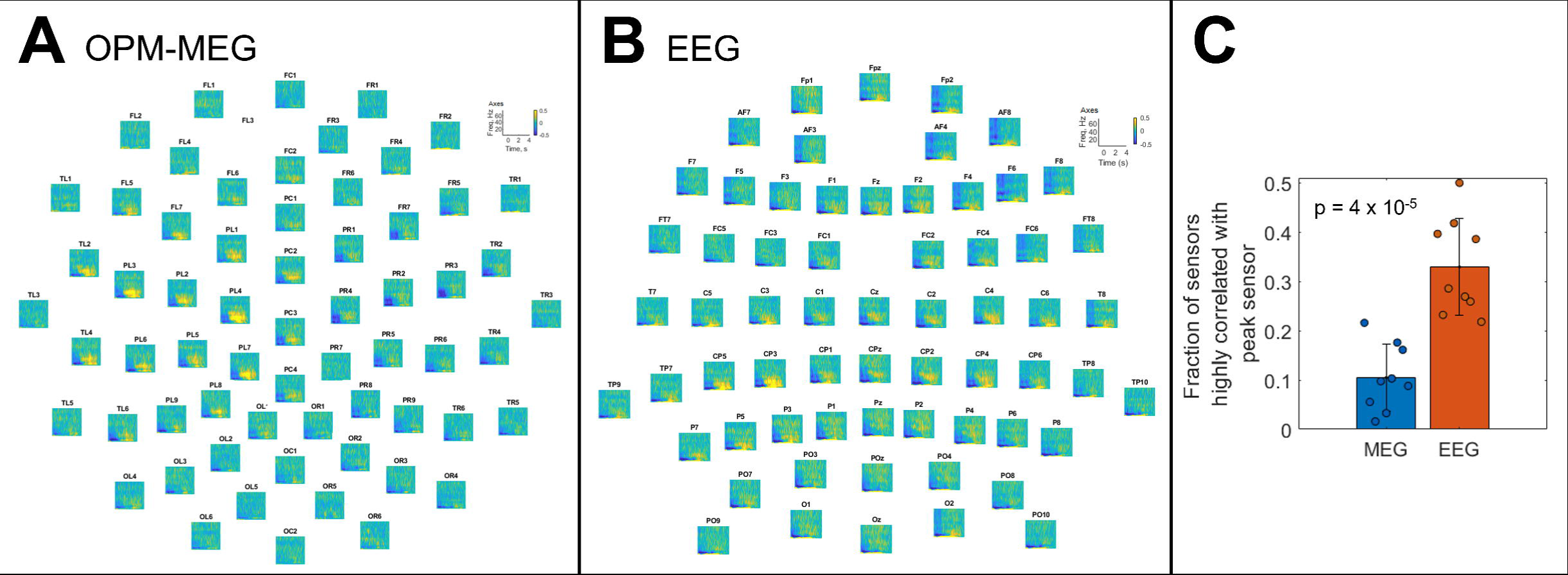
Volume conduction and field spread: Panels A and B show the time-frequency spectrograms across all sensors, for simultaneous OPM-MEG (left) and EEG (right) in a single representative participant. The EEG has been re-referenced to the average of all sensors. There is visibly greater signal spread across the scalp in EEG compared with OPM-MEG, as would be expected. Signal spread is quantified in panel C where the fraction of sensors that are highly correlated (R>0.3) with the peak sensor is plotted. Data points show individuals, the bars represent the mean value, and error bars represent standard deviation. The fraction of highly correlated sensors is significantly lower in OPM-MEG compared with EEG.

### Alpha generation task

Table 2 shows the number of participants, trials and channels remaining in each dataset after preprocessing.

Figure 5 shows, in a single representative participant, the increase in alpha activity when the eyes are closed. The time courses in Figures 5A and 5B show simultaneously acquired EEG and OPM-MEG recordings over a 10 s period (5-s eyes closed; 5-s eyes open). Alpha oscillations are clear in both modalities and are reduced when the eyes are opened. A spectral analysis for this participant is shown in Figures 5C/D, for all channels in EEG (panel C), and the radial channels in OPM-MEG (panel D). PSDs are plotted for the eyes open segments (blue) and eyes closed segments (orange) with a clear peak at ∼10Hz in the posterior channels, when eyes were closed.

**Figure 5:**
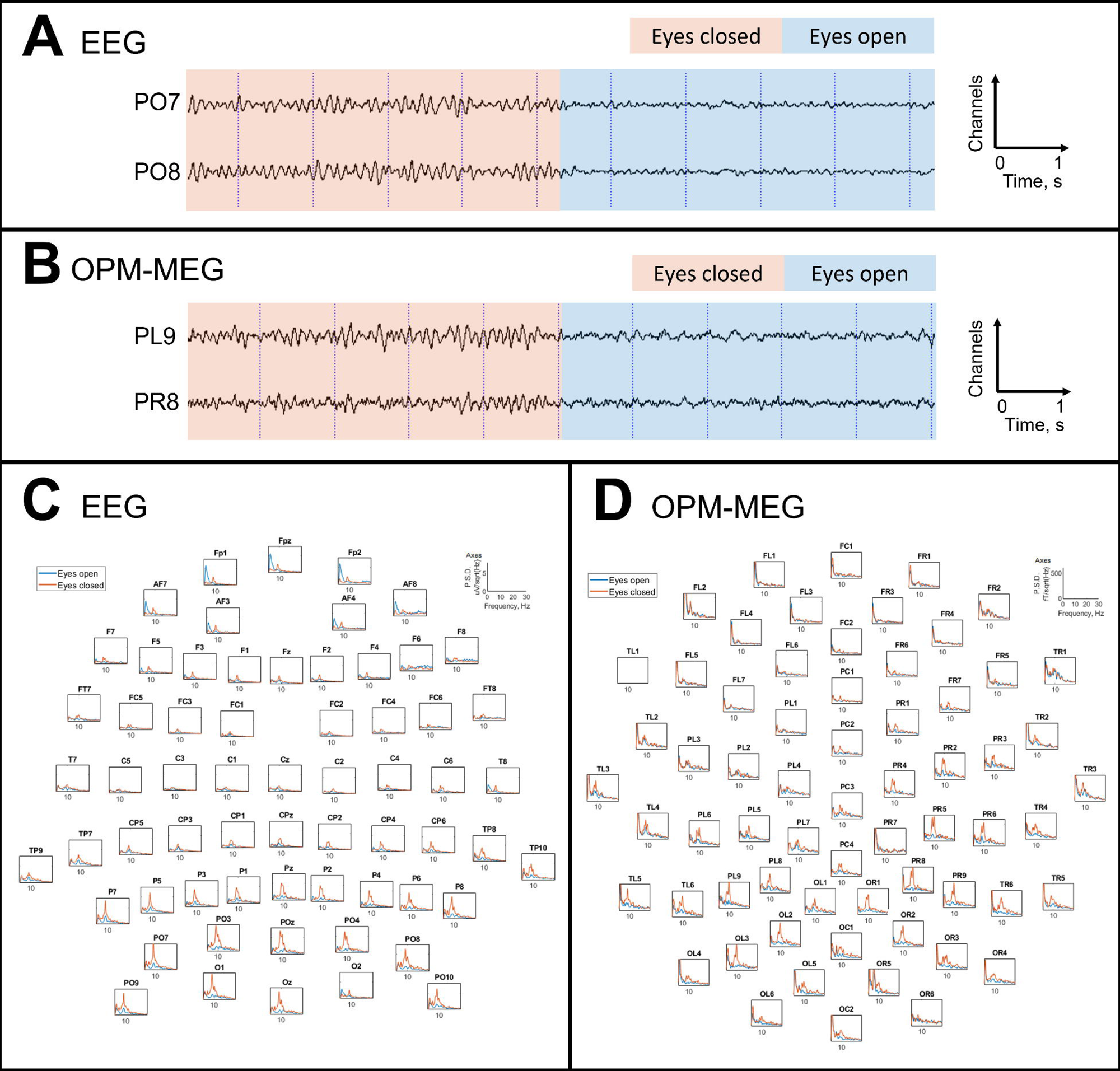
Alpha activity generated by eye closure in a representative participant, for simultaneous EEG and OPM-MEG. Panels A and B show simultaneous EEG and OPM-MEG data, filtered between 2 and 40Hz. Panels C and D show the PSD plots for each channel across the whole head for EEG and OPM-MEG respectively. There is a clear alpha peak (∼10Hz) during the eyes closed period which dominates posterior channels, as expected.

The channels with the highest signal contrast (between eyes open and closed) were extracted for all participants and the participant-averaged PSDs are shown in Figure 6; eyes open in blue, eyes closed in orange. Figures 6A and 6B show EEG only and EEG recorded in the presence of OPM-MEG. Figures 6D and 6E show OPM-MEG only and OPM-MEG recorded in the presence of EEG. Again, the presence of the additional modality appears to have little effect on the signal contrast. Figures 6C and 6F formalise this finding by showing signal contrast values for EEG (C) and OPM-MEG (F). A Wilcoxon rank-sum test suggested that OPM-MEG has no effect on signal contrast in EEG (p = 0.9) and likewise that EEG has no effect on signal contrast in OPM-MEG (p = 0.98). This, in agreement with the results in Figure 3, suggests that concurrent OPM-MEG/EEG is feasible.

**Figure 6:**
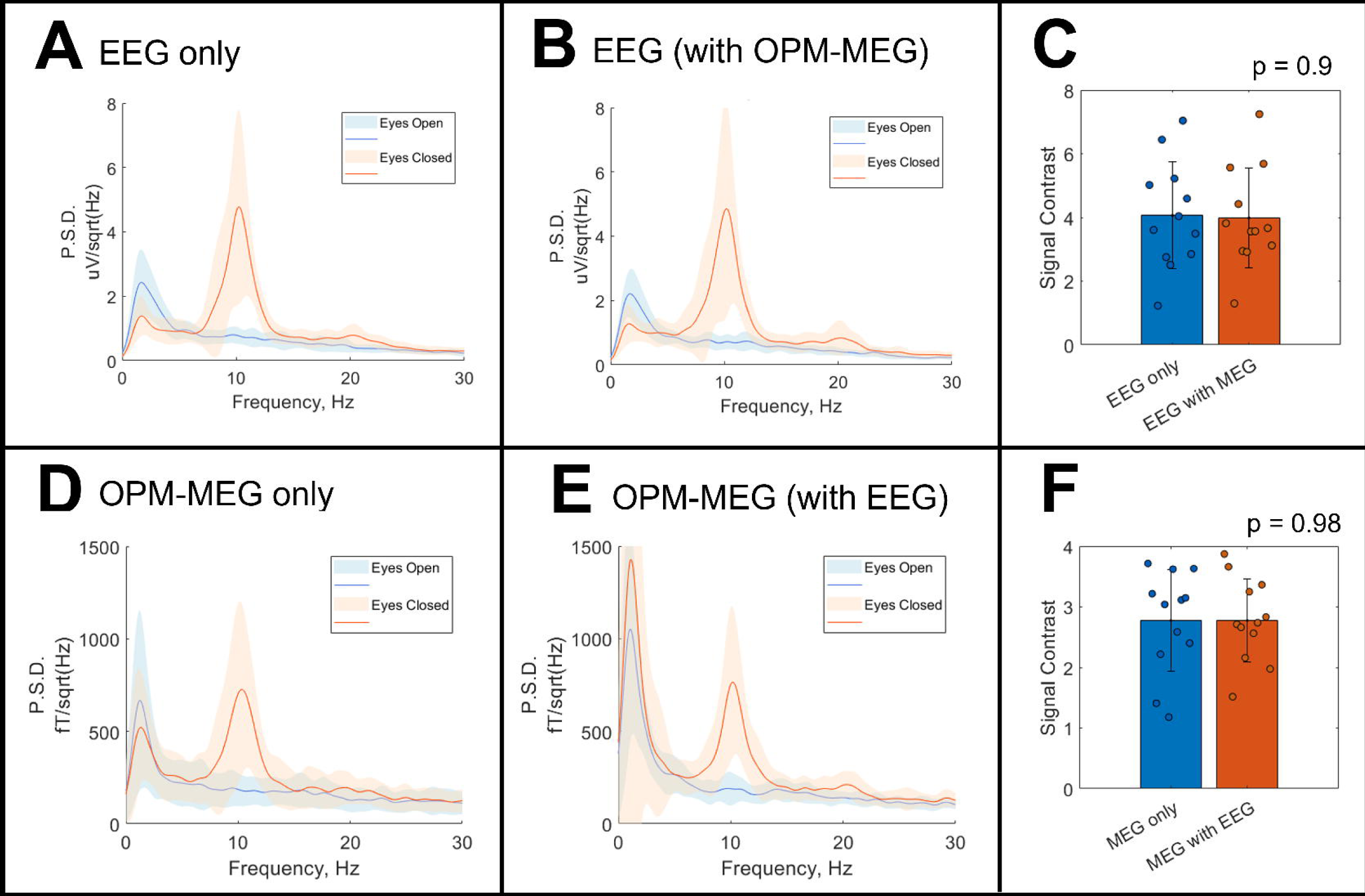
Spectral analysis of the alpha generation task. Panels A and B show the PSD plots from the best channels for EEG alone and EEG in the presence of OPM-MEG respectively. The solid line is the mean over participants, and the shaded area represents standard deviation. The EEG signal contrast values are shown in panel C: data points represent single individuals; the bar shows the mean and the error bar standard deviation. Panels D and E show the PSDs for OPM-MEG alone and in the presence of EEG respectively. Panel F shows the signal contrast values.

## DISCUSSION

The aim of this study was to show that simultaneous, wearable, whole-head EEG and OPM-MEG is feasible. To this end we have acquired data across two tasks: a motor task known to modulate beta oscillations, and an eyes-open / eyes-closed task known to modulate alpha oscillations. In the former, the SNR of the beta response was similar, regardless of whether modalities were used individually or concurrently. Likewise in the latter experiment, signal contrast was stable regardless of the concurrent recording. Importantly, the amplitude of the background magnetic field inside the room (a critical consideration to enable free participant movement in OPM-MEG) was also unchanged by the presence of EEG. These results suggest that simultaneous wearable EEG/OPM-MEG is possible.

The utility of simultaneous EEG and conventional MEG has been well documented: the high sensitivity to tangential sources in MEG, and the high sensitivity to radial sources in EEG, are complementary and from a theoretical point of view this improves uniformity of coverage and sensitivity. Moreover, from a clinical perspective, multiple studies have demonstrated that simultaneous recordings offer non-redundant information, and this is reflected in clinical practice guidelines (set out by the American Clinical Magnetoencephalography Society (ACMEGS)) which state that standard scalp EEG should be recorded simultaneously with MEG for assessment of patients with epilepsy (Bagic, Knowlton, Rose, & Ebersole, 2011). This is not only advantageous due to non-redundant information but is also useful to grow confidence in MEG: EEG has been used for many years for the diagnosis and management of disorders including epilepsy, tumours, dementia, head injury (including concussion) and encephalitis, whereas the high cost and poor practicality of conventional MEG have led to an under-utilisation (Bagić, et al., 2023). Concurrent recording means that clinicians who are less familiar with MEG can keep the widely established EEG measures whilst simultaneously gaining new information from MEG. The usability of a wearable platform, coupled with the high spatial resolution and sensitivity afforded by OPM-MEG (compared to conventional MEG) (Boto, et al., 2019) (Hill, et al., 2020) (Rhodes, et al., 2023) mean that concurrent OPM-MEG and EEG is even more attractive than the fusion of EEG with conventional MEG. For these reasons, the findings presented in this paper are important.

Although there was no measurable difference in SNR between OPM-MEG data acquired with and without EEG, there is a small reduction in mean OPM-MEG SNR, when simultaneous EEG was added (See Figure 3F) and this warrants consideration. For the measurements acquired we used two independent systems and so the presence of the EEG electrodes necessarily moved the OPMs slightly further from the scalp. Since the MEG signal amplitude drops with the square of the distance from the source, even the relatively low thickness of an EEG electrode and cap (∼ 3 mm) will move the OPMs sufficiently to reduce signal strength noticeably. This was masked by inter-individual differences in the present data, and so the drop in amplitude was not significant. Nevertheless, the presence of EEG will (marginally) reduce the amplitude of the OPM-MEG signal and, for this reason, future systems might aim to build EEG electrodes directly in to the OPM-MEG helmet. This should be possible, particularly if new generations of OPM-MEG system involve flexible (EEG-cap-like) helmets.

An important result in this paper is the finding in Figure 4 that the proportion of “highly correlated” sensors was larger in EEG than in OPM-MEG. In EEG, electrical potentials are “smeared out” across the scalp by a combination of volume conduction, and the high resistivity of the skull. The former means that a single source in the brain affects the signal at multiple sensors, whereas the latter makes this process hard to mathematically model (Baillet, 2017). The result is that, as shown in Figure 4B, the signal (in this case from motor cortex) is spread to a large number of channels and this, coupled with the difficulty in modelling, leads to EEG having limited spatial resolution. In MEG, the magnetic field propagates from a single source to multiple sensors, resulting in a similar problem (though the relative transparency of the skull to magnetic field makes the MEG forward problem more tractable, and consequently gives MEG a better spatial resolution). The extent of the field spread depends on proximity of the sensors to the scalp and leads to more diffuse signals in conventional MEG (where cryogenic sensors are necessarily more distal) than OPM-MEG (where sensors are closer to the scalp surface). It is known that the ability to disentangle two sources in the brain depends strongly on how much the fields correlate at the sensor level (Boto, et al., 2019) (Brookes, et al., 2021) (Hill, et al., In Submission), and therefore the more the field spreads (i.e. the higher the proportion of highly correlated sensors across the scalp) the lower the ultimate spatial resolution of the technique will be. Our simple analysis therefore demonstrates directly the significant advantage of OPM-MEG over EEG in terms of spatial specificity – we observe a significantly more focal field pattern.

There are some limitations of the present study that should not be overlooked. Firstly, our findings relate only to two experimental paradigms, modulation of beta oscillations by finger movement and modulation of alpha oscillations by opening and closing the eyes. There are other signals that could have been measured (e.g., evoked responses). It seems likely that the present findings will extend to these other signals, however this may not necessarily be the case (e.g., for high frequencies, EEG signals are obfuscated by fields from muscle activity (Muthukumaraswamy, 2013) (Boto, et al., 2019)). Thus, future studies should aim to characterise fully the similarities and differences between EEG and OPM-MEG data acquired simultaneously across the whole frequency spectrum from delta through to high gamma bands. Second, a key limitation of EEG is that it measures the difference in electric potential between each scalp electrode and a reference, meaning results can change depending on where the reference is chosen. For the results presented here we chose an average reference so that the EEG was not reliant on a single scalp electrode. To ensure that our results were robust, we repeated the analyses with the common recording reference (CRR, electrode FCz) and results are shown in supplementary information. We found that the impact of changing the referencing system was minimal, nevertheless this choice of reference is likely to affect further signal analyses (e.g., source localisation). For our signal spread analyses, the number of channels was different for EEG (63 excluding ECG) and OPM-MEG (128), and MEG channels were recorded in two different orientations (radial and tangential with respect to the scalp). This makes a direct comparison between modalities difficult, as does the distance between adjacent sensors. However, the distance between adjacent EEG and OPM-MEG sensors was similar (∼4cm) with roughly even coverage of 63 and 64 sensors across the scalp respectively. Finally, here we have deliberately only employed channel-space analysis; we do not propose that this is the best way to present MEG data, rather we made this choice to enable a direct testing of our hypotheses and to avoid confounds associated with the relative advantages and disadvantages of source reconstruction methodologies. Had we chosen to undertake source reconstruction, this would likely improve the signal to noise of both OPM-MEG and EEG, but the fundamental finding (that both EEG and OPM-MEG work irrespective of the other modality) would remain.

## CONCLUSION

In this study we successfully acquired simultaneous whole-head EEG and OPM-MEG data in 12 healthy adults. Our results show that there is no statistically significant difference in OPM-MEG signal quality with or without the presence of EEG and vice versa. This indicates that future clinical or research studies can employ simultaneous EEG and OPM-MEG (with appropriate MEG-compatible equipment) without significantly affecting data quality.

## AUTHOR CONTRIBUTIONS

**ZAS**: Conceptualisation, Methodology, Software, Formal analysis, Investigation, Data Curation, Writing – Original Draft, Writing – Review and Editing, Visualisation, Project Administration; **KStP**: Conceptualisation, Methodology, Investigation, Resources, Writing – review and Editing; **NH**: Methodology, Software, Writing – Review and Editing; **MR**: Methodology, Software; **LA**: Conceptualisation, Investigation; **TMT**: Methodology, Software, Resources, Writing – Review and Editing; **RP**: Resources; **KJM**: Conceptualisation, Methodology, Writing – Review and Editing; **JHC**: Conceptualisation, Supervision, Writing – Review and Editing; **EB**: Conceptualisation, Methodology; **MJB**: Conceptualisation, Methodology, Resources, Writing – Original Draft, Writing – Review and Editing, Supervision, Funding Acquisition.

## Data Availability

The data and code used in this study are available on GitHub: https://github.com/ZSeedat/EEG_OPMMEG_HealthyAdultStudy_YoungEpilepsy

## ACKNOWLEDGEMENTS

This work was supported by an Engineering and Physical Sciences Research Council (EPSRC) Healthcare Impact Partnership Grant (EP/V047264/1) and a Biomedical catalyst grant funded by Innovate UK (10037425). We acknowledge support from the UK Quantum Technology Hub in Sensing and Timing, funded by EPSRC (EP/T001046/1). The OPM-MEG system was purchased with funding from the Wolfson Foundation. JHC’s research is made possible by the NIHR Great Ormond Street Hospital Biomedical Research Centre. TT is funded by an ERUK and Young Epilepsy emerging leader award (FY2101) and by the National Brain Appeal Innovation Fund.

