## Supplementary Information for "Simultaneous whole-head electrophysiological recordings using EEG and OPM-MEG"

The nature of the EEG signal means that it is recorded relative to a reference electrode. Different montages can be used to view the EEG recording in different ways – for example, referencing to an electrode at the front of the head will amplify signals at the back of the head. In our analyses in the main manuscript, we employed an average reference – in other words, each electrode was measuring activity relative to the average over all electrodes (excluding noisy channels). This makes the EEG data more comparable with MEG which measures an absolute value of magnetic field (without reference to a magnetic field at another location). To make sure that the results were not dependent on the referencing scheme employed, analyses were also repeated with the common reference. The following figures (S1 – S4) show those results.

For the motor induced response task, the results for the peak channels remain largely unchanged (Figure S1). In other words, the task modulated response (motor related beta desynchronisation (MRBD) followed by a post-movement beta rebound (PMBR)) is clearly visible in both EEG and OPM-MEG measured alone or simultaneously and there is no significant difference in signal-to-noise ratios (SNRs) when simultaneous recordings took place. The difference between reference schemes is minimal: for EEG alone, the SNR was 10 ± 5 (mean ± std) for the average reference, and 9 ± 3 (mean ± std) for the CRR. For EEG in the presence of OPM-MEG the SNR was 9 ± 6 (mean ± std) when either the average reference or the CRR was used.

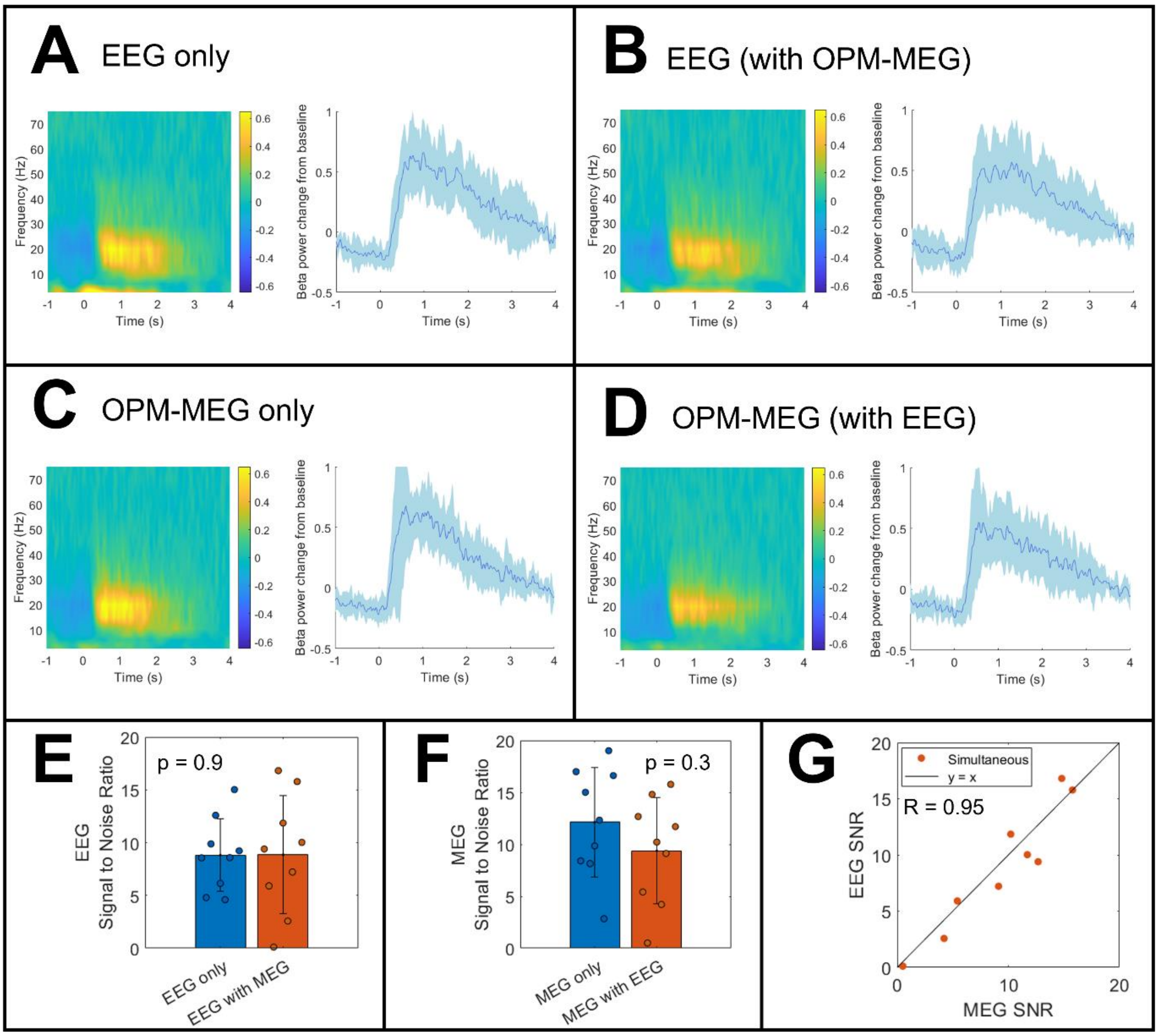


*Figure S1: Right index finger abduction results for the peak channel in both EEG and OPM-MEG data. For EEG the CRR was used. Time-frequency spectrograms and beta amplitude time courses are shown for EEG alone (A), EEG in the presence of OPM-MEG (B), OPM-MEG alone (C) and OPM-MEG in the presence of EEG (D). All four results show a clear task-modulated response in the beta band (MRBD followed by PMBR). Panels E and F show the SNRs of EEG and OPM-MEG respectively. Error bars show the standard deviation. There was no significant difference in EEG SNR when OPM-MEG was present (p = 0.86), similarly the was no significant difference in OPM-MEG SNR when EEG was present (p = 0.34). Panel G illustrates that the variation in SNR across participants is likely down to their individual brain differences – people exhibiting a low SNR in MEG recordings also had a low SNR in EEG.*

However, the choice of referencing scheme had much greater impact on our measure of signal spread (Figure S2). The task-modulated response in EEG appears to be more diffuse when the CRR is used so that the fraction of sensors highly correlated with the peak sensor changes from 0.5 ± 0.3 (mean ± std) for the average reference to 0.8 ± 0.1 (mean ± std) for the CRR. In either case, there is a significantly higher proportion of sensors highly correlated to the peak sensor in EEG than in OPM-MEG as the theory predicts.

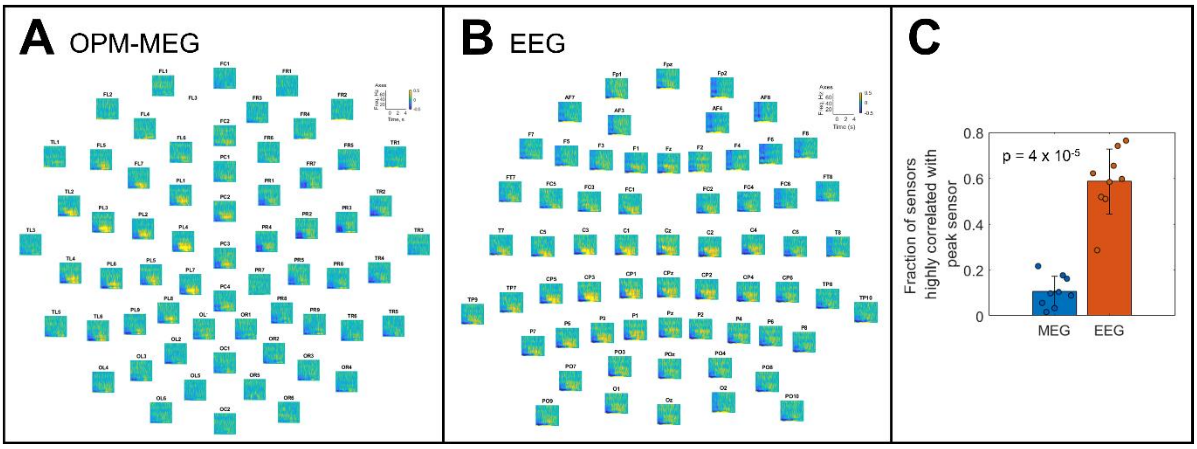


*Figure S2: Signal spread during the right index finger abduction task; for EEG the CRR was used. Panels A and B show the trial-averaged time-frequency spectrograms across the whole head in an example participant for simultaneous OPM-MEG and EEG results respectively. Panel C which shows the fraction of channels highly correlated (R >0.6) with the peak channel. EEG has significantly more highly correlated channels than OPM-MEG (p = 5x10$^{-4}$).*

For the alpha generation task, traces were again visibly inspected but with the CRR for EEG. Figure S3 shows these results for an example participant. In both EEG and OPM-MEG there is a visible alpha rhythm (panels A and B respectively). Frequency analysis across all channels shows a clear peak at approximately 10Hz, maximal posteriorly for both EEG and OPM-MEG in agreement with our initial results. It is worth noting that there is markedly less alpha visible in EEG frontal channels with this choice of reference in comparison with the average reference (even when posterior channels had been excluded from the average), see Figure S3 and Figure 4 in the main manuscript.

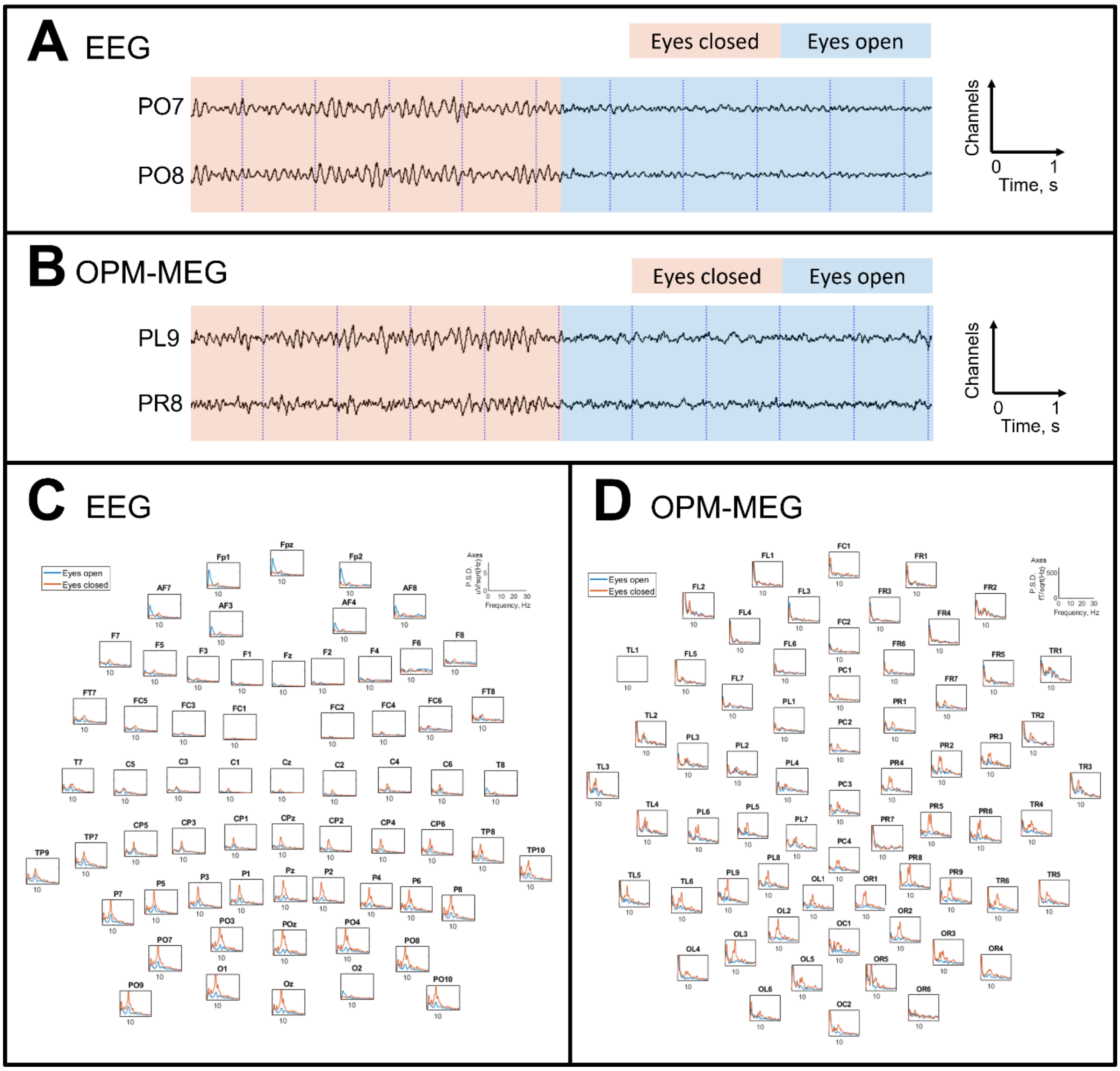


*Figure S3: Alpha activity generated by eye closure in an example participant. The CRR was used for EEG. Panels A and B show an excerpt of the EEG and OPM-MEG signals respectively (bandpass filtered 2-40Hz) for a simultaneous recording. A clear alpha rhythm is visible which abolishes on eye opening. Power spectral density plots across the whole head are shown for EEG (C) and OPM-MEG (D). There is a clear peak at approx. 10Hz in posterior channels during eyes closed segments for both modalities.*

For each participant the peak sensor was selected, and power spectral density (PSD) plots were computed and then averaged across all participants. Figure S4 shows these PSDs for both EEG and OPM-MEG, with the common recording reference used for EEG. In all 4 cases (EEG alone, EEG recorded with OPM-MEG, OPM-MEG alone, and OPM-MEG recorded with EEG) there is a prominent alpha peak during the eyes closed period. Panels C and F show the signal contrast between eyes open and eyes closed conditions for EEG and OPM-MEG respectively. In agreement with our initial findings, there is no significant difference in signal contrast between EEG alone (3 ± 1) (mean ± std) and EEG in the presence of OPM-MEG (4 ± 1) (mean ± std), p = 0.8.

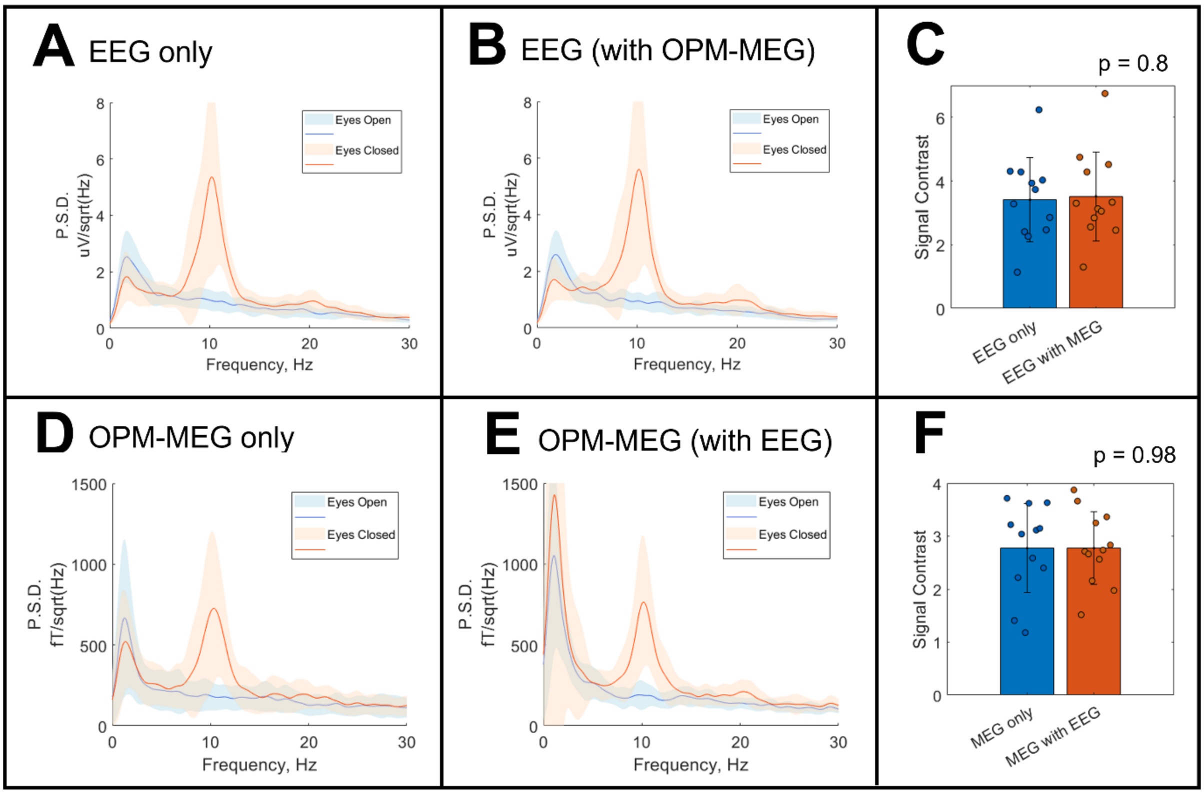


*Figure S4: Frequency decomposition of the peak channels averaged across participants for EEG alone (A), EEG in the presence of OPM-MEG (B), OPM-MEG alone (D), and OPM-MEG in the presence of EEG (E). The solid line is the mean, and the shaded area shows the standard deviation across participants. In each case there is a prominent peak at approx. 10Hz during the eyes closed condition (orange) compared with eyes open (blue). Panels C and F show the signal contrast for EEG and OPM-MEG respectively. The bars represent the mean, error bars represent the standard deviation, and the scatter points are individual participants. There is no significant difference between the individual and simultaneously acquired recordings for either EEG (p = 0.8) or MEG (p = 0.98).*
